# Lingering sex and age disparities in dolutegravir uptake among adults with HIV: A multi-country observational cohort study

**DOI:** 10.1101/2025.05.23.25325682

**Authors:** Ellen Brazier, Matthew L Romo, Andrea Ciaranello, Francesca Odhiambo, Sanjay Pujari, Gad Murenzi, Charles Kasozi, Sasisopin Kiertiburanakul, Dominique Mahambou Nsonde, Winnie R. Muyindike, Vohith Khol, Patricia Lelo, Rita Lyamuya, Man Po Lee, Denis Nash, IeDEA

## Abstract

**Introduction:** Since July 2019, the World Health Organization (WHO) has recommended dolutegravir (DTG)-based regimens as preferred first-line antiretroviral therapy (ART) for adults and adolescents living with HIV (DTG-for-All), a reversal of a 2018 safety alert on use of DTG-based regimens by women of reproductive age (WRA). We examined sex and age disparities in DTG uptake before and after DTG-for-All in the International epidemiology Databases to Evaluate AIDS (IeDEA).

**Methods:** We included patients ≥16 years on or initiating treatment between January 2017 and July 2021 in 14 low- and middle-income countries where initial guidelines on DTG-based regimens for first-line ART either restricted use by WRA or had no such restrictions. We estimated the cumulative incidence of DTG uptake (CI-DTG) by sex and age group (aged 16-49 years vs. 50+ years), stratified by patient, clinic and setting characteristics.

**Results:** Among 177,706 patients on ART during the study period, 51% were females aged 16-49 years, with 25% males aged 16-49, and 13% and 11%, respectively, females and males aged 50+. At the time of DTG-for-All, overall CI-DTG was 29.6% (95%CI: 29.4, 29.8); it was lower among females aged 16-49 (16.2%; 95%CI: 16.0-16.5) than males (41.1%; 95%CI: 40.6, 41.5), with no sex disparities among patients aged 50+ (females: 46.0%; males: 47.0%). While DTG uptake subsequently increased among all groups, by July 2021, it remained substantially lower among females 16-49 (66.4%; 95%CI: 66.1, 66.7), compared with males 16-49 and older females and males (75.8% to 77.5%). Concentrated in countries where initial guidelines on DTG restricted use by WRA, disparities in DTG uptake persisted at all health system levels and in both low-income and lower-middle income countries.

**Conclusions:** While sex-age differentials in DTG uptake narrowed after WHO’s DTG-for-All recommendation, lingering disparities uptake underscore the challenges of policy de-implementation when new evidence emerges.

**Key Messages:** *What is already known on this topic:* Prior research illuminated substantial sex and age group disparities in uptake of dolutegravir (DTG)-based antiretroviral therapy (ART) regimens among people with HIV (PLHIV)—disparities that emerged after the World Health Organization (WHO) issued a May 2018 drug safety alert for women of reproductive age (WRA) and persisted after subsequent guidance in July 2019 recommending DTG-based regimens as preferred first-line treatment for all adults and adolescents, irrespective of age and sex.

*What this study adds:* Drawing on real-world service delivery data from 72 clinics in 14 countries, this study documents the persistence of sex and age group disparities in DTG-based regimen uptake. Concentrated in low- and lower-middle income countries where national guidelines initially contained restrictions on use of DTG by WRA, these disparities remained substantial at two and three years after WHO’s “dolutegravir for all” guidance among treatment-naïve and treatment-experienced PLHIV and were observed at all health system levels.

*How this study might affect research, practice or policy:* Our study underscores the substantial lags that can occur in updating, disseminating, and translating new policy guidance into practice, as well as the challenges of de-implementing outdated policy recommendations when new evidence emerges—challenges that will remain relevant with new treatment regimens, such as injectable ART, on the horizon.

## Introduction

Since mid-2019, the World Health Organization (WHO) has recommended DTG-based regimens for all adults and adolescents (DTG-for-All), including women and girls irrespective of age and contraceptive use [1]. This recommendation reversed WHO’s May 2018 drug safety alert [2], issued in response to an interim analysis of surveillance data from the Tsepamo cohort in Botswana showing higher-than-expected incidence of neural tube defects (NTDs) among infants born to women on DTG at the time of conception [3]. With additional Tsepamo data showing a diminished association between DTG and NTDs [4], along with modeling studies examining population-level risks and benefits that supported DTG use among women of reproductive age (WRA) [5, 6], WHO’s 2019 treatment guidelines unconditionally recommended DTG as first-line treatment. In many countries, however, public health agencies had restricted access to DTG-based regimens for WRA in response to WHO’s 2018 safety alert [7, 8], and these restrictions continued after the DTG-for-All recommendation. An early analysis [9] of DTG uptake in 11 low and middle-income countries in the International epidemiology Databases to Evaluate AIDS (IeDEA) research collaboration showed that large differences by sex and age emerged after WHO’s 2018 safety alert and persisted after WHO’s 2019 DTG-for-All recommendation, with only 29% of females aged 16-49 initiating a DTG-based regimen in early 2020, compared with 58% of males in the same age group.

Given extensive data showing the superior efficacy and tolerability of DTG and its potential population health benefits compared with older regimens [10], sustained disparities in DTG access and uptake could negatively impact HIV care outcomes among WRA. Using updated data from many of the same IeDEA clinics and cohorts as our previous study [9], this analysis aimed to examine whether sex and age disparities in DTG uptake persisted at two and three years after WHO’s DTG-for-All recommendation and to examine how trends in DTG uptake differed by clinic, setting, and patient characteristics.

## Methods

### Data sources

This retrospective cohort study used data from the IeDEA research collaboration [11], which pools de-identified routine clinical data on more than 2 million people with HIV (PWH) ever enrolling in HIV care at care and treatment sites in 44 countries [12]. Our study population was drawn from IeDEA cohorts in the Asia-Pacific, Central Africa, and East Africa regions. Our study observation period was from January 2017 through July 2021 (i.e., two years after DTG-for-All) and through July 2022 (three years after DTG-for-All) for a subset clinics with sufficient follow-up time before database closure.

For each country in our study, we identified and reviewed national guidelines on use of DTG as first-line antiretroviral therapy (ART), based on UNAIDS policy data for 2017-2022 [13] and local HIV treatment guidelines [14–26]. We categorized countries as having restrictive guidelines for DTG use if initial guidelines on DTG-based regimens as first-line therapy restricted use by WRA or included requirements for contraceptive use and/or informed consent by WRA. We categorized countries as having non-restrictive guidelines for DTG use if guidelines initially recommended DTG-based regimens as the preferred first-line for all adults and adolescents, irrespective of sex, age and pregnancy, or guidelines recommended DTG but offered an alternative (e.g., efavirenz) for WRA who “opted out” of DTG-based regimens after counseling.

For our analysis of DTG uptake two years after DTG-for-All, clinics were included if pharmacy records reflected any use of DTG-based regimens and if the database closure date was after July 31, 2021. Clinics were excluded if they had fewer than 50 patients in care or if less than one percent of patients were initiated on a DTG-based regimen over the entire study observation period. Patients were eligible if they had any record of ART; were at least 16 years of age at the time of initiating treatment; and had at least one visit after the date that their site of care began prescribing DTG-based regimens. Patients were excluded for the following reasons: HIV status recorded as negative; sex unknown or missing, implausible dates of death (e.g., before dates of treatment or other clinical contacts); implausible dates of initiating a DTG-based regimen (relative to other patients at the same clinic); or incomplete or missing ART regimen information.

A subset of sites with a database closure date after July 31, 2022 were included in our analysis of DTG uptake three years after DTG-for-All, with the same patient eligibility and exclusion criteria applied for these sites.

### Measures

The primary study outcome was DTG uptake, which was defined as initiating a DTG-based regimen among ART-naïve patients or switching to a DTG-based regimen among patients already on ART.

Our main exposure measures were sex, as recorded in medical records (male or female), and age group. Age was calculated at the time of either ART start or at the time the patient’s clinic started prescribing DTG-based regimens, whichever came later, and was categorized into two groups (16 to 49 years and 50+ years) to align with the period typically considered reproductive age for women (15-49 years) [27].

Other patient characteristics of interest were history of ART use at the time their clinic began prescribing DTG-based regimens (ART-naïve vs. experienced) and ART anchor drug of current regimens: DTG, efavirenz, nevirapine, protease inhibitors (PIs), other integrase strand transfer inhibitors (INSTIs), or other non-nucleoside reverse transcriptase inhibitors (NNRTIs). Clinic and contextual characteristics included health facility type (health center, district hospital, or regional, provincial or university hospital); urban or rural location; restrictive or non-restrictive country guidelines on DTG for first-line ART (as described above); and World Bank classification of country income level (low-income, lower-middle income, or upper-middle income) at the time of clinic rollout of DTG [28].

### Statistical analysis

We used descriptive statistics to summarize sample characteristics, overall and by sex and age group, as well as to characterize uptake of DTG-based regimens among ART-naïve and ART-experienced patients; previous regimen bases for patients switched to DTG-based regimens; and latest regimen bases among patients alive and in care at the time database closure. Among patients in care and not on DTG-based regimens at database closure, we examined current ART regimen base, by sex and age group.

We used competing risks regression with the Aalen–Johansen estimator [29] to estimate crude cumulative incidence proportions (i.e., risk) of DTG uptake (CI-DTG) with 95% confidence intervals (95%CIs) at four timepoints: the WHO safety alert (i.e., May 18 2018), the DTG-for-All recommendation (July 22 2019), and one and two years after DTG-for-All (July 31 2020 and July 31 2021, respectively), as described in our previous work [9]. For clinics with a database closure date after July 31, 2022, we estimated crude cumulative incidence proportions of DTG uptake at an additional timepoint three years after DTG-for-All (i.e., July 31, 2022). Cumulative incidence proportions were estimated and plotted for each sex and age group and further stratified by patient and clinic characteristics of interest, including ART experience (i.e., naïve vs. not naïve at the start of follow-up); restrictive vs. non-restrictive country guidelines on DTG for first-line ART; country income level, and health facility type. We also computed and plotted the ratio of female to male cumulative incidence proportions at each timepoint to visualize how sex and age group disparities changed over time.

Because patients enrolled in care in Kenya comprised almost half of our dataset, in sensitivity analyses, we assessed whether overall sex and age group disparities in CI-DTG at two years after DTG-for-All were similar when Kenya sites were excluded.

All statistical analyses were performed using SAS 9.4 (SAS Institute, Cary, NC).

### Ethical Approval

Data were approved for use by local research ethics committees in each participating IeDEA region, as well as the institutional review boards of their data management centers.

### Patient and Public Involvement

We used de-identified routinely collected data, and there was no contact with patients. Accordingly, patient and public involvement in the design, conduct, reporting or dissemination of our research was not applicable.

## Results

### Sample characteristics

Among 179,807 eligible patients at clinics with a database closure date after 31 July 2021, 2,101 (1.2%) were excluded because of missing data related to sex, implausible death dates, incomplete ART medication data, or negative HIV status (Supplementary Fig 1), resulting in a sample of 177,706 patients across 72 clinics in 14 countries: Burundi (3.6%), Cambodia (0.3%), Cameroon (7.5%), Congo (3.5%), Democratic Republic of Congo (DRC) (2.0%), India (0.9%), Indonesia (0.1%), Kenya (48.4%), Malaysia (0.3%), Rwanda (9.7%), Tanzania (5.4%), Thailand (0.4%), Uganda (17.9%), and Vietnam (0.2%) (Table 1).

**Table 1.** Sample of patients at sites with a database closure after July 31, 2021 (72 clinics in 14 countries)

| **Characteristic** | **All Patients** | **Females**<br>**Aged 16-49 y** | **Males**<br>**Aged 16-49 y** | **Females**<br>**Aged ≥50 y** | **Males**<br>**Aged ≥50 y** |
| --- | --- | --- | --- | --- | --- |
| Total, n (%) | 177706 | 90939 (51.2) | 44477 (25.0) | 23467 (13.2) | 18823 (10.6) |
| Mean age (SD), y | 41.6 (11.3) | 36 (7.9) | 38.3 (7.2) | 56.7 (5.9) | 57.2 (6.2) |
| **Experience with ART at start of follow-up, n (%)** |  |  |  |  |  |
| Experienced | 135326 (76.2) | 67658 (74.4) | 31062 (69.8) | 20549 (87.6) | 16057 (85.3) |
| Naïve | 42380 (23.8) | 23281 (25.6) | 13415 (30.2) | 2918 (12.4) | 2766 (14.7) |
| **Site urbanicity, n (%) \*** |  |  |  |  |  |
| Rural | 36112 (20.3) | 18501 (20.3) | 8757 (19.7) | 5233 (22.3) | 3621 (19.2) |
| Urban | 141594 (79.7) | 72438 (79.7) | 35720 (80.3) | 18234 (77.7) | 15202 (80.8) |
| **Site level of care, n (%) \*** |  |  |  |  |  |
| Health centers | 48869 (27.5) | 26208 (28.8) | 11931 (26.8) | 6154 (26.2) | 4576 (24.3) |
| District hospitals | 43124 (24.3) | 22650 (24.9) | 10502 (23.6) | 5867 (25.0) | 4105 (21.8) |
| Regional, university or referral hospitals | 85713 (48.2) | 42081 (46.3) | 22044 (49.6) | 11446 (48.8) | 10142 (53.9) |
| **Country, n (%)** |  |  |  |  |  |
| Burundi | 6373 (3.6) | 2888 (3.2) | 1086 (2.4) | 1352 (5.8) | 1047 (5.6) |
| Cambodia | 500 (0.3) | 175 (0.2) | 175 (0.4) | 61 (0.3) | 89 (0.5) |
| Cameroon | 13345 (7.5) | 6717 (7.4) | 3164 (7.1) | 2147 (9.1) | 1317 (7.0) |
| Congo | 6132 (3.5) | 2993 (3.3) | 732 (1.6) | 1430 (6.1) | 977 (5.2) |
| Democratic Republic of Congo | 3497 (2.0) | 1987 (2.2) | 589 (1.3) | 536 (2.3) | 385 (2.0) |
| India | 1516 (0.9) | 448 (0.5) | 628 (1.4) | 118 (0.5) | 322 (1.7) |
| Indonesia | 235 (0.1) | 54 (0.1) | 115 (0.3) | 20 (0.1) | 46 (0.2) |
| Kenya | 86014 (48.4) | 44869 (49.3) | 20873 (46.9) | 11602 (49.4) | 8670 (46.1) |
| Malaysia | 448 (0.3) | 50 (0.1) | 246 (0.6) | 32 (0.1) | 120 (0.6) |
| Rwanda | 17163 (9.7) | 8618 (9.5) | 5110 (11.5) | 1741 (7.4) | 1694 (9.0) |
| Tanzania | 9590 (5.4) | 5134 (5.6) | 2199 (4.9) | 1302 (5.5) | 955 (5.1) |
| Thailand | 700 (0.4) | 181 (0.2) | 297 (0.7) | 85 (0.4) | 137 (0.7) |
| Uganda | 31888 (17.9) | 16714 (18.4) | 9109 (20.5) | 3030 (12.9) | 3035 (16.1) |
| Vietnam | 305 (0.2) | 111 (0.1) | 154 (0.3) | 11 (0) | 29 (0.2) |
| **Initial guidelines on DTG-based regimens for 1st-line ART, n (%)** |  |  |  |  |  |
| Restrictive guidelines for WRA† | 159015 (89.5%) | 82022 (90.2%) | 40221 (90.4%) | 20489 (87.3%) | 16283 (86.5%) |
| Non-restrictive guidelines for WRA‡ | 18691 (10.5%) | 8917 (9.8%) | 4256 (9.6%) | 2978 (12.7%) | 2540 (13.5%) |
| **Country income, n (%)** |  |  |  |  |  |
| Low income | 58921 (33.2) | 30207 (33.2) | 15894 (35.7) | 6659 (28.4) | 6161 (32.7) |
| Lower-middle income | 117637 (66.2) | 60501 (66.5) | 28040 (63.0) | 16691 (71.1) | 12405 (65.9) |
| Upper-middle income | 1148 (0.6) | 231 (0.3) | 543 (1.2) | 117 (0.5) | 257 (1.4) |
| **IeDEA region, n (%)** |  |  |  |  |  |
| Central Africa | 46510 (26.2) | 23203 (25.5) | 10681 (24.0) | 7206 (30.7) | 5420 (28.8) |
| East Africa | 127492 (71.7) | 66717 (73.4) | 32181 (72.4) | 15934 (67.9) | 12660 (67.3) |
| Asia-Pacific | 3704 (2.1) | 1019 (1.1) | 1615 (3.6) | 327 (1.4) | 743 (3.9) |

ART: Antiretroviral therapy. DTG: Dolutegravir. SD: Standard deviation. WRA: Women of reproductive age.
† Burundi, Cambodia, Cameroon, Democratic Republic of Congo, Indonesia, Kenya, Rwanda, Uganda
‡ Congo, India, Malaysia, Tanzania, Thailand, Vietnam

Overall, females aged 16-49 years comprised 51.2% of the sample, with 25.0% males aged 16-49, and 13.2% and 10.6%, respectively, females and males aged 50 years and above. Mean age at ART start was 41.6 years (standard deviation 11.3), and most (79.2%) were ART-experienced at the start of follow-up. Most patients (79.7%) were from clinics in urban areas, with little variation by patient age and sex. Almost half (48.2%) received HIV care at regional, provincial or university hospitals, with 27.5% in care at health centers and 24.3% at district hospitals. The overwhelming majority of patients were in low-income (33.2%) and lower-middle income (66.2%) countries in IeDEA’s Central and East Africa cohorts (97.9%), with only 0.6% from upper-middle income countries in IeDEA’s Asia-Pacific region. Most patients (89.5%) were in countries where initial guidelines recommending DTG as the preferred first-line treatment—issued between 2018 and 2021—were restrictive, as defined in Methods (Supplementary Table 1); 10.5% of patients were in countries where initial guidelines on DTG for first-line treatment were not restrictive.

### Use of DTG-based regimens

Among 135,326 patients already on ART at the time their clinic began prescribing DTG-based regimens, 94,074 (69.5%) were switched to a DTG-based regimen during the study observation period, ranging from 64.9% among females aged 16-49 to 72.6% among males aged 16-49, 76.3% among females aged

50+ and 74.0% among males aged 50+ (Table 2). Among 42,380 ART-naïve patients initiating treatment after their clinic began prescribing DTG-based regimens (Supplementary Table 2), 56.8% were first initiated on a DTG-based regimen, ranging from 45.7% of females aged 16-49 to 70.7% among males aged 16-49, 68.2% of females aged 50+, and 70.2% of males aged 50+. Among those initiated on non-DTG-based regimens during the study observation period, half (50.0%) later switched to a DTG-based regimen, ranging from 48.2% of females aged 16-49 to 51.6% of males aged 16-49, 61.6% of females aged 50+, and 56.9% of males aged 50+.

**Table 2.** Cumulative Incidence of Dolutegravir Uptake, by Sex and Age Group.

| Patient characteristic | Summary of Outcomes |  |  |  | Dolutegravir Uptake, Cumulative Incidence Proportion (95% CI) |  |  |  |
| --- | --- | --- | --- | --- | --- | --- | --- | --- |
|  | Total Patients, *n* | Patients Ever Initiating DTG, *n* | Patients Who Dropped Out, *n* | Patients Censored, *n* | Before the Safety Signal (Until 18 May 2018) | After the Safety Signal (Until 22 July 2019) | One year after WHO Recommended Dolutegravir for All (Until July 22 2020) | Two years after WHO Recommended Dolutegravir for All (Until July 22 2021) |
| **Entire sample** | **177706** | **127292** | **24888** | **25526** | **2.8 (2.7, 2.9)** | **29.6 (29.4, 29.8)** | **48.0 (47.7, 48.2)** | **71.4 (71.2, 71.6)** |
| **Sex and age group** |  |  |  |  |  |  |  |  |
| Females aged 16-49 y | 90939 | 60648 | 15023 | 15268 | 2.7 (2.5, 2.8) | 16.2 (16.0, 16.5) | 33.6 (33.3, 33.9) | 66.4 (66.1, 66.7) |
| Males aged 16-49 y | 23467 | 18236 | 2213 | 3018 | 2.3 (2.2, 2.4) | 41.1 (40.6, 41.5) | 61.8 (61.4, 62.3) | 76.5 (76.1, 76.8) |
| Females aged ≥50 y | 44477 | 34107 | 5671 | 4699 | 3.7 (3.5, 4.0) | 46.0 (45.4, 46.7) | 64.1 (63.5, 64.7) | 77.5 (77.0, 78.1) |
| Males aged ≥50 y | 18823 | 14301 | 1981 | 2541 | 3.4 (3.2, 3.7) | 47.0 (46.2, 47.7) | 64.4 (63.8, 65.1) | 75.8 (75.2, 76.4) |
| **Experience with ART at start of follow-up** |  |  |  |  |  |  |  |  |
| **Experienced** | **135326** | **94074** | **18053** | **23199** | **3.3 (3.2, 3.4)** | **33.8 (33.6, 34.1)** | **49.8 (49.5, 50.0)** | **69.4 (69.1, 69.6)** |
| Females aged 16-49 y | 67658 | 43914 | 10197 | 13547 | 3.2 (3.0, 3.3) | 19.2 (18.9, 19.5) | 34.8 (34.5, 35.2) | 64.7 (64.3, 65.1) |
| Males aged 16-49 y | 31062 | 22596 | 4173 | 4293 | 2.7 (2.6, 2.9) | 47.3 (46.8, 47.9) | 63.7 (63.2, 64.3) | 72.6 (72.1, 73.1) |
| Females aged ≥50 y | 20549 | 15674 | 1956 | 2919 | 4.1 (3.8, 4.4) | 48.9 (48.2, 49.5) | 65.4 (64.7, 66.0) | 76.1 (75.5, 76.7) |
| Males aged ≥50 y | 16057 | 11890 | 1727 | 2440 | 3.8 (3.5, 4.1) | 50.2 (49.4, 50.9) | 65.7 (64.9, 66.4) | 74.0 (73.3, 74.6) |
| **Naïve** | **42380** | **33218** | **6835** | **2327** | **1.2 (1.1, 1.3)** | **16.2 (15.8, 16.5)** | **42.3 (41.8, 42.7)** | **77.8 (77.4, 78.2)** |
| Females aged 16-49 y | 23281 | 16734 | 4826 | 1721 | 1.1 (1.0, 1.3) | 7.5 (7.1, 7.8) | 30.1 (29.5, 30.7) | 71.3 (70.7, 71.8) |
| Males aged 16-49 y | 13415 | 11511 | 1498 | 406 | 1.3 (1.1, 1.5) | 26.6 (25.8, 27.3) | 57.4 (56.6, 58.3) | 85.3 (84.7, 85.9) |
| Females aged ≥50 y | 2918 | 2562 | 257 | 99 | 1.1 (0.7, 1.5) | 26.2 (24.6, 27.8) | 55.4 (53.6, 57.2) | 87.4 (86.1, 88.5) |
| Males aged ≥50 y | 2766 | 2411 | 254 | 101 | 1.6 (1.1, 2.1) | 28.3 (26.6, 30.0) | 57.3 (55.4, 59.1) | 86.5 (85.2, 87.7) |

Among 134,284 patients in care at the time of database closure, 31,080 (23.1%) were not on DTG-based regimens, of whom 18,317 (58.9%) were females aged 16-49; 5,736 (18.5%) males aged 16-49; 3,864 (12.4%) females 50+ years; and 3,163 (10.2%) males 50+years (Supplementary Table 2). Among these patients, females aged 16-49 were more likely to be on Efavirenz-based regimens (60.8%), compared with men aged 16-49 (46.7%) or older females and males (40.1% and 39.1%, respectively), and they were less likely than other age groups to be on PI- or other INSTI-based regimens (32% vs. 43-47%) (Supplementary Fig 2).

### Cumulative incidence of Dolutegravir uptake and differences by sex and age group

By the time of WHO’s safety alert in May 2018, CI-DTG—either newly initiating or switching—was 2.8% (95% CI: 2.7, 2.9), with little substantive difference by sex and age group (Table 2). However, large disparities emerged after WHO’s safety alert, and by the time of WHO’s DTG-for-All recommendation (July 2019), CI-DTG was 16.2% (95%CI: 16.0, 16.5) among females aged 16-49, compared with 41.1% (95%CI: 40.6, 41.5) among males aged 16-49; 47.0% (95%CI: 46.2, 47.7) among males aged 50+; and 46.0% (95%CI: 45.4, 46.7) among females aged 50+. Sex and age group disparities attenuated over time; however, CI-DTG remained substantively lower among WRA at two years after DTG-for-All (66.4%; 95%CI: 66.1, 66.7) compared with males aged 16-49 (76.5%, 95% CI: 76.1, 76.8), males aged 50+ (75.8%, 95% CI: 75.2, 76.4) and older females (77.5%, 95% CI: 77.0, 78.1). Sex and age group disparities in CI-DTG were observed among ART-naïve patients initiating treatment, as well as ART-experienced patients switching to DTG-based regimens. Among females aged 16-49, CI-DTG at two years after DTG-for-All increased monotonically for each 10-year age group, from 61.2% (95%CI: 60.5, 61.8) among 20–29-year-olds to 64.3% (95%CI: 63.8, 64.8) and 71.9% (95%CI: 71.4-72.3), respectively, among 30–39-year-olds and 40–49-year-olds (Fig 1).

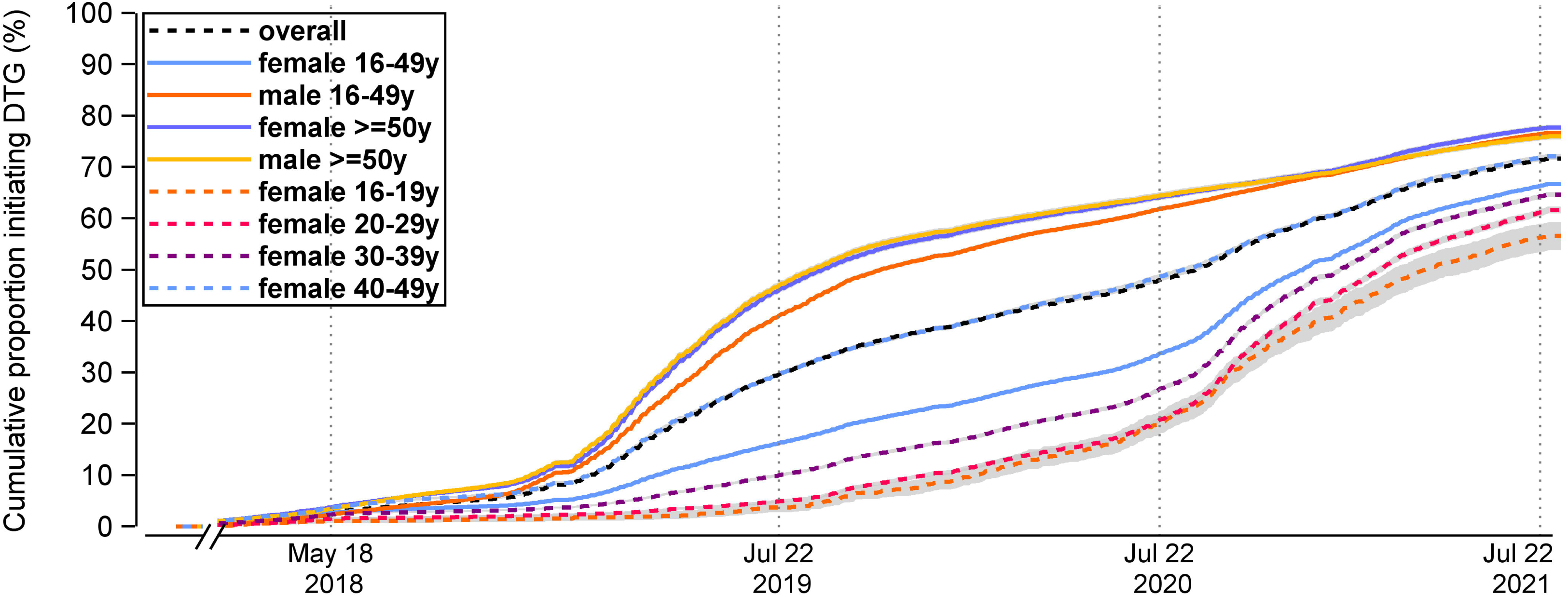

In sensitivity analyses that excluded clinics in Kenya, overall CI-DTG and sex and age group disparities in DTG uptake were consistent with our main analysis (Supplementary Table 3).

### CI-DTG uptake and differences by contextual and clinic characteristics

Table 3 presents CI-DTG by sex and age group, stratified by country and clinic characteristics. Two years after DTG-for-All, overall CI-DTG was similar in countries where clinical guidelines on DTG initially included restrictions for WRA (71.7%; 95%CI: 71.5, 72.0) and countries with non-restrictive guidelines (68.2%; 95%CI: (67.5, 68.9). However, large sex and age group disparities were present two years after DTG-for-All in countries with restrictive guidelines, with no such disparities in countries with non-restrictive guidelines. In countries with restrictive guidelines, sex and age group disparities in DTG uptake were notable among patients initiating treatment (Fig 2, Panel A) and among ART-experienced patients switching to DTG-based regimens (Panel B)—gaps that remained substantial two years after DTG-for-All. In contrast, in countries with non-restrictive guidelines, there were no sex and age group differences in DTG initiation (Panel C) or switching (Panel D), and in these settings, females aged 16-49 were more likely to be switched to DTG-based regimens than males or older females.

**Table 3.**
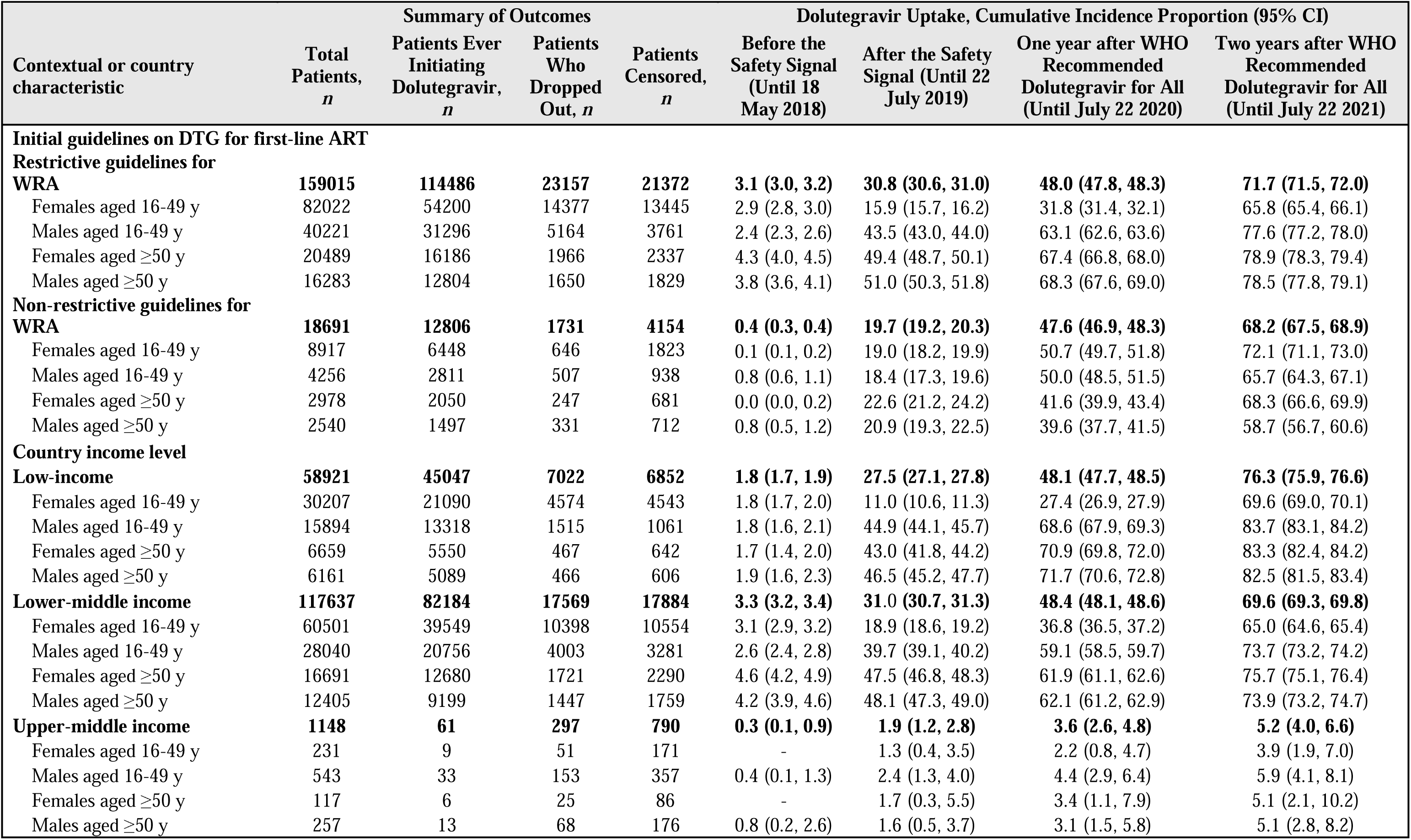
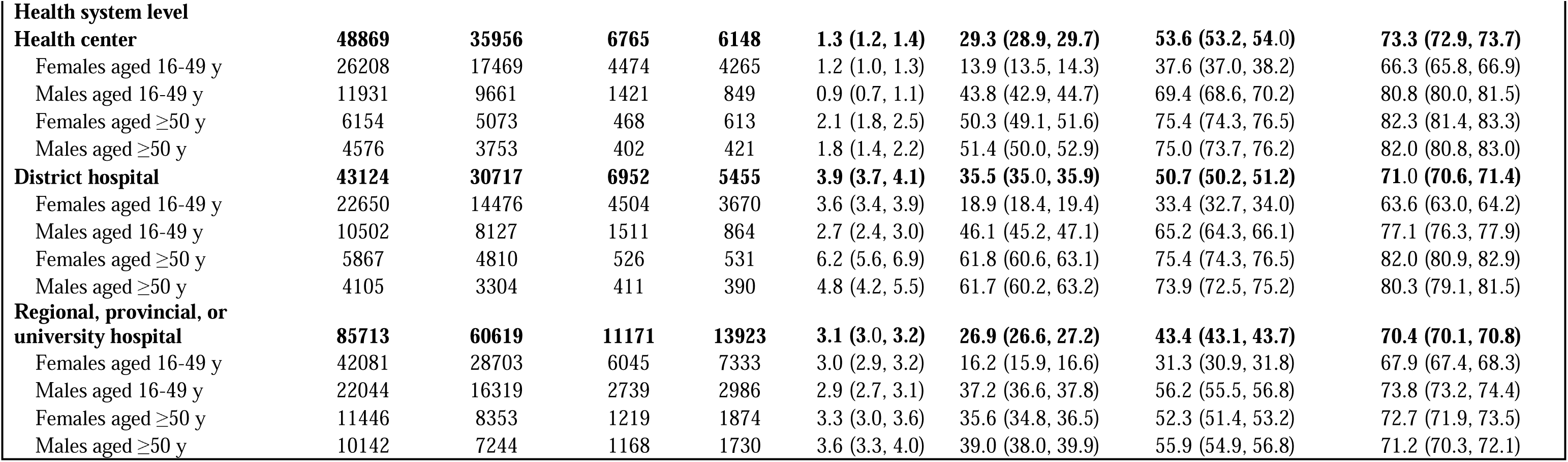
Age- and Sex-Group Specific Cumulative Incidence of Dolutegravir Uptake, by contextual characteristic.

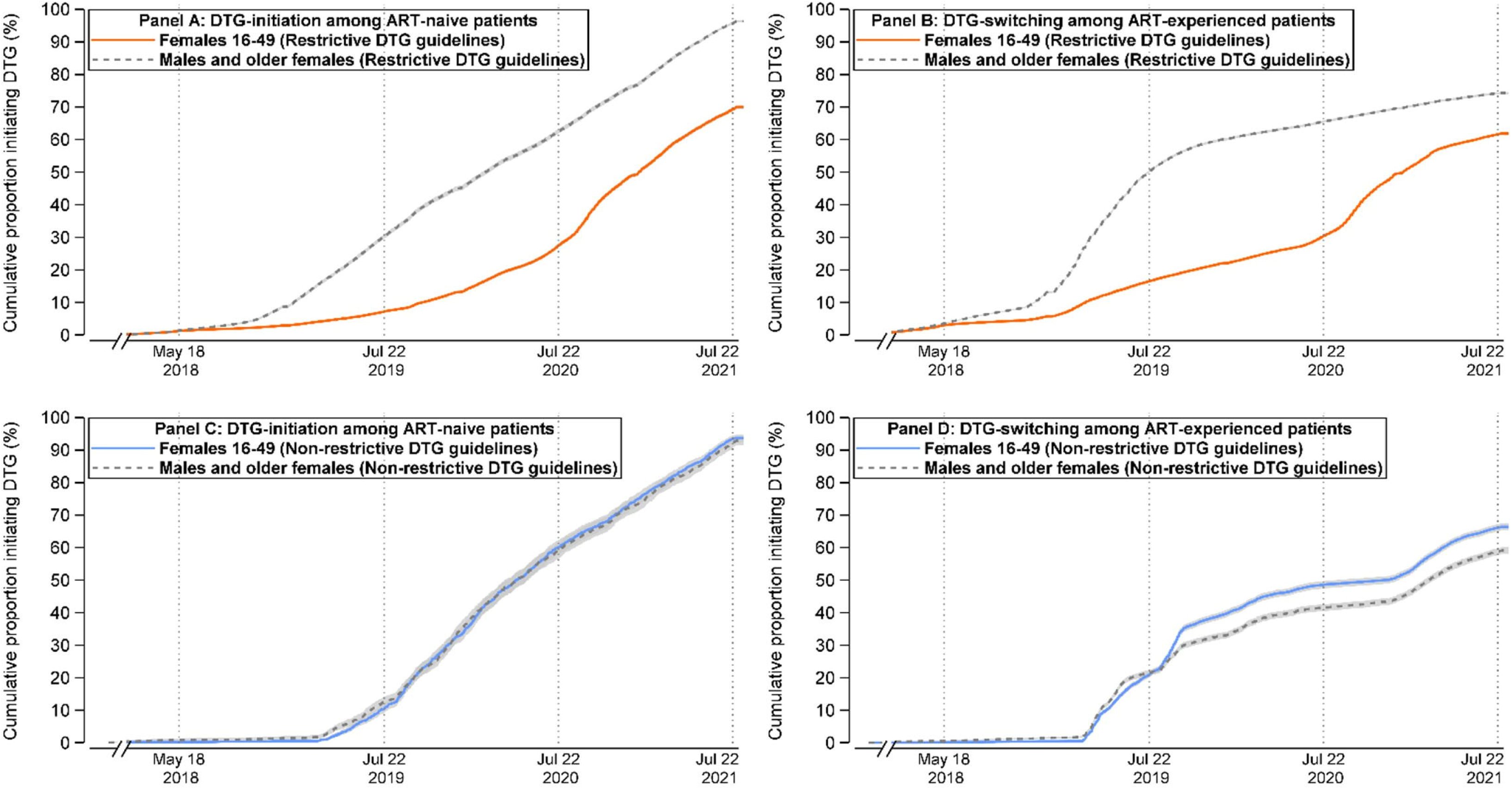

Two years after DTG-for-All, overall CI-DTG was highest in low-income (76.3%; 95% CI: 75.9, 76.6) and lower-middle income countries (69.6%; 95% CI: 69.3, 69.8), and it remained substantially lower in upper-middle-income countries (5.2%; 95% CI: 4.0, 6.6) (Table 3). Absolute sex and age group disparities in DTG uptake were also greatest in low-income and lower-middle countries and were negligible in upper-middle income countries. Overall CI-DTG two years after DTG-for-All did not differ substantially across levels of the health system (Health centers: 73.3%; 95%CI: 72.9, 73.7; District hospitals: 71%; 95%CI: 70.6, 71.4; Regional, provincial or university hospitals: 70.4%; 95%CI: 70.1, 70.8). However, sex and age group disparities in DTG uptake were observed at all facility types.

### DTG uptake by three years after WHO’s DTG-for-All recommendation

Among 185,159 eligible patients at clinics with a database closure date after July 31, 2022, similar proportions (1.3%) were excluded because of missing data related to sex, implausible death dates, incomplete ART medication data, or non-negative HIV status, resulting in a sample of 182,824 patients across 66 clinics in eight countries: Burundi (3.6%), Cameroon (7.6%), Congo (3.4%), Democratic Republic of Congo (DRC) (2.2%), Kenya (49.5%), Rwanda (10.0%), Tanzania (5.7%), and Uganda (18.0%). Patient characteristics for the sample of clinics with a database closure date after July 2022 were similar to those of patients at clinics with an earlier database closure date (Supplementary Table 4).

While overall CI-DTG reached 78.7% (95%CI: 78.5, 78.9) three years after DTG-for-All, sex and age group disparities persisted, with CI-DTG at 75.7% (95%CI: 75.4, 75.9) among females aged 16-49, compared with 81.6% to 82.2% among males and older females (Supplementary Table 5). Sex and age group disparities in DTG uptake by July 2022 were observed among both ART-naïve and ART-experienced patients, as well as across low-income and lower-middle income settings, and all levels of the health system (Supplementary Table 6). Consistent with our main analysis, these disparities were observed in countries with initially restrictive guidelines on DTG for first-line ART and were not observed in countries where initial guidance on DTG did not include restrictions for WRA.

## Discussion

Our study of 72 clinics across 14 countries found that sex and age disparities in uptake of DTG-based ART regimens that emerged after WHO’s 2018 safety alert narrowed, but remained substantial, a full two years after WHO’s DTG-for-All recommendation, with uptake of DTG-based regimens 15% higher among males and older females than females aged 16-49. Additionally, in eight countries where follow-up time before database closure allowed us to examine trends at three years after DTG-for-All, DTG uptake was 8% higher among males and older females than females aged 16-49.

Disparities in DTG uptake were concentrated in countries where national guidelines on use of DTG for first-line ART initially included restrictions on use of DTG by WRA. Most of these countries subsequently issued updated guidelines during the study period, removing restrictions on DTG-based regimens for WRA. Nonetheless, lingering disparities persisted two and three years after WHO’s DTG-for-All recommendation. In contrast, in countries where clinical guidelines introducing DTG-based regimens for first-line ART did not include restrictions or cautions about use by WRA, sex and age disparities in DTG uptake did not emerge. Our findings reinforce conclusions of our earlier research that initial restrictions on DTG use by WRA could have an enduring effect, despite new evidence and updated WHO guidance [9]. They also accord with findings from other recent research [30, 31], showing substantial sex and age group disparities in DTG uptake after WHO’s DTG-for-All recommendation, both between and within countries.

With its parallels to ongoing denial and limited uptake of menopausal hormonal therapy by women who might benefit from it [32], lingering disparities in DTG uptake in countries with initial restrictions for use by WRA underscore the challenge of policy de-implementation—namely the difficulty of stopping practices and interventions that are no longer evidence based [33, 34]. Notably, three countries in our study issued guidelines restricting use of DTG-based regimens by WRA after WHO’s DTG-for-All recommendation, and updating of restrictive guidelines lagged WHO’s recommendations by one to three years despite support from PEPFAR for the development of national guidelines to advance the transition to DTG-based regimens [35–38].

High uptake of DTG-based ART by men of all ages and older females in low and lower-middle income countries suggest that medication stock-outs were not a barrier to DTG uptake ART by WRA in these settings. Instead, our findings suggest that policy-, clinic-, provider- and patient-level factors continued to constrain use of DTG-based regimens by WRA after WHO recommended DTG unconditionally. Of note, we found similar patterns across all types of health facilities, with disparities even observed at regional, provincial and university teaching hospitals, which would be expected to implement new guidelines more quickly. We also observed that among females aged 16-49, DTG uptake decreased with each ten-year decrease in age, with those in their teens and twenties least likely to be initiated on or transitioned to DTG-based regimens. These results suggest that those most likely to become pregnant may continue to be discouraged from using DTG-based regimens, despite evidence of its safety, and they underscore the need for guideline changes to be reinforced with provider training, as well as awareness-raising efforts for PLWH to ensure that patients are informed about all treatment options available.

While sex and age group disparities were not observed in upper-middle income countries in IeDEA’s Asia-Pacific region, we observed very low uptake of DTG in all age and sex groups in these settings. These findings raise questions about why patients were not accessing DTG. Updates to national guidelines recommending DTG as the preferred first-line treatment and national procurement of DTG occurred later in this region than in sub-Saharan Africa [13, 39], and a survey of IeDEA sites in 2020 found that 33% of clinics in the Asia-Pacific region (33%) did not use DTG for first- or second-line ART, compared with 0% of clinics in Central and East Africa [40]. As most Asia-Pacific countries reflected in our study were covered by the Medicines Patent Pool (MPP) and ViiV Healthcare’s 2014 licensing agreement [41], which allowed generic manufacturers to produce low-cost versions of DTG for low- and middle-income countries, supply- and demand-side barriers to the uptake of DTG-based ART in upper-middle income countries merit further research.

Several study limitations are worth noting. We have limited data on patient characteristics and preferences, and it is possible that disparities in DTG uptake reflect enduring patient preferences based on outdated information about the safety of DTG-based regimens for WRA. We also have limited data on the reasons for regimen switching, and it is difficult to distinguish between first- or second-line use of DTG-based regimens in many data systems. While it is possible that higher CI-DTG switching among males reflects greater need for second-line regimens among this group, rather than restricted access for WRA, this is unlikely to explain much of the lingering sex and age group disparities in DTG uptake observed in our study.

A further limitation is that we lack data on HIV care providers’ knowledge and training, and sites participating in IeDEA are not nationally representative in many settings, particularly in countries where IeDEA sites are hospital-level clinics in urban centers. However, such sites generally have better access to up-to-date information on standards of care, and they usually have the capacity to provide more advanced levels of care than peripheral health facilities. Accordingly, the magnitude of sex and age-based disparities may be underestimated in some countries in our study. Given the difficulty of accessing historical HIV treatment guidelines and circulars, and it is also possible that some countries published earlier recommendations on DTG-based ART than those which we identified.

These limitations notwithstanding, an important strength of our study is our large dataset reflecting real-world service delivery at a diverse range of countries and clinics, and our ability to examine DTG uptake two and three years after WHO’s DTG-for-All recommendation. While prior research [32] examined DTG uptake in some of the same countries, the study sample was more limited, reflecting patients enrolled in care at 12 clinics across four counties. Drawing on data from more than 70 clinics across all levels of the health system in 14 countries, our results provide additional evidence that non-negligible disparities in uptake of DTG-based regimens persist and that additional efforts may be needed to address outdated provider and patient perceptions about DTG safety in order to fully realize the potential population-level benefits of DTG. Our findings underscore the substantial lags that can occur in updating, disseminating, and translating new policy guidance into practice, as well as the challenges of de-implementing outdated policy recommendations when new evidence emerges—challenges that will remain relevant with new ART regimens, such as injectable ART on the horizon.

## Data Availability

Data Availability Statement: Complete data for this study cannot be posted in a supplemental file or a public repository because of legal and ethical restrictions. The principles of collaboration of IeDEA and the regulatory requirements of the different IRBs of our participating sites (sometimes representing national IRBs of ministries of health) require the submission of a project concept proposal and approval by the IeDEA Executive Committee. To request data, please review IeDEA guidance available at: https://www.iedea.org/resources/multiregional-research-sops-templates/ and contact the Executive Committee (https://www.iedea.org/working-groups/executive-committee/). Signing of a data sharing agreement may also be required.

## Acknowledgements

The authors acknowledge the patients and staff at all facilities who contributed data to this study, as well as the site investigators and data managers across the IeDEA collaboration.

## Author contributions

**Conceptualization**: Matthew L. Romo, Denis Nash, Ellen Brazier,

**Data curation**: Francesca Odhiambo, Sanjay Pujari, Gad Murenzi, Charles Kasozi, Sasisopin Kiertiburanakul, Dominique Mahambou Nsonde, Winnie R. Muyindike, Vohith Khol, Patricia Lelo, Rita Lyamuya, Man Po Lee.

**Funding acquisition**: Denis Nash.

**Data analysis**: Ellen Brazier.

**Visualizations**: Ellen Brazier.

**Writing – original draft**: Ellen Brazier.

**Writing – review & editing**: Matthew Romo, Andrea Ciaranello, Denis Nash, Francesca Odhiambo, Sanjay Pujari, Gad Murenzi, Charles Kasozi, Sasisopin Kiertiburanakul, Dominique Mahambou Nsonde, Winnie R. Muyindike, Vohith Khol, Patricia Lelo, Rita Lyamuya, Man Po Lee.

## Funding acknowledgement

The International Epidemiology Databases to Evaluate AIDS (IeDEA) is supported by the U.S. National Institutes of Health’s National Institute of Allergy and Infectious Diseases, the *Eunice Kennedy Shriver* National Institute of Child Health and Human Development, the National Cancer Institute, the National Institute of Mental Health, the National Institute on Drug Abuse, the National Heart, Lung, and Blood Institute, the National Institute on Alcohol Abuse and Alcoholism, the National Institute of Diabetes and Digestive and Kidney Diseases, and the Fogarty International Center: Asia-Pacific, U01AI069907; Central Africa, U01AI096299; and East Africa, U01AI069911. Informatics resources are supported by the Harmonist project, R24AI24872. This work is solely the responsibility of the authors and does not necessarily represent the official views of any of the institutions mentioned above.

