## Supplementary materials for "Lingering sex and age disparities in dolutegravir uptake among adults with HIV: A multi-country observational cohort study"

**Supplementary Figure 1: Flow diagram for study participants**

*Patients with >30 days between recorded date of ART initiation and the earliest ART medication recorded in pharmacy data and patients with unknown ART regimens.

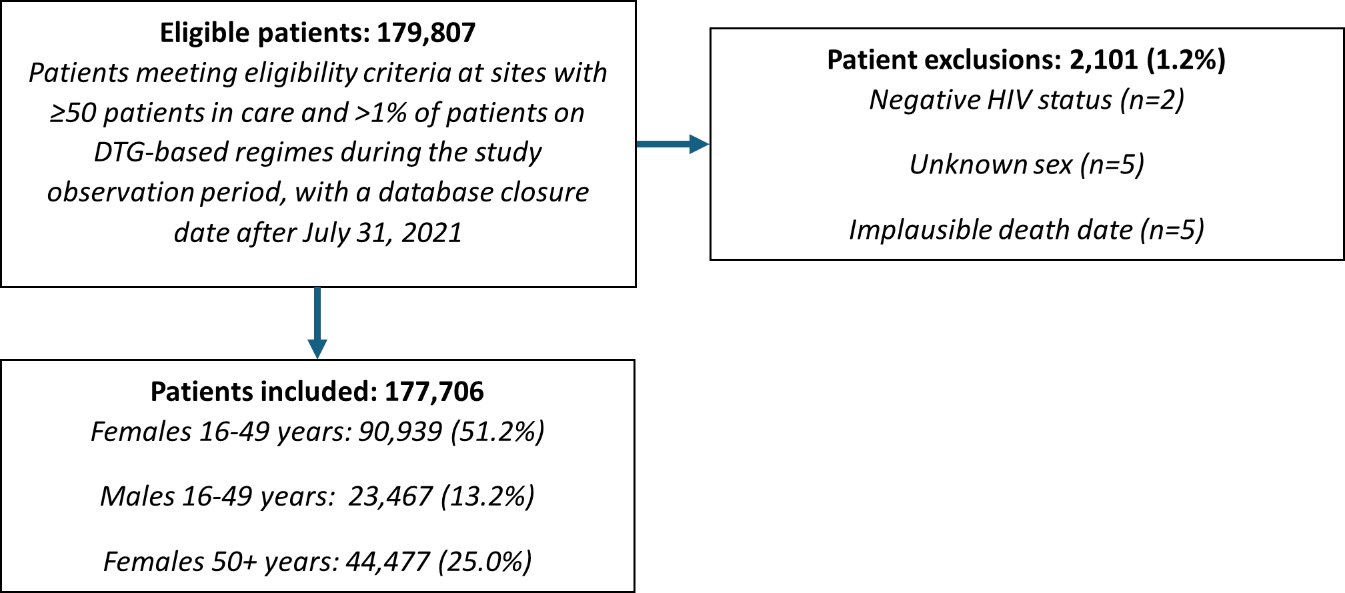

**Supplementary Table 1. National guidelines on DTG for first-line ART**

| **Country** | **Initial Policy Guidelines on DTG (Year)** | **Guidance** | **Restrictive policy (for WRA)** | **Unconditional DTG recommendation (Year)** |
| --- | --- | --- | --- | --- |
| Burundi | Addendum aux directives nationales d’ultisation des ARVs (2018) [1] | *Translation*: “In 2017, WHO recommended the introduction of Dolutegravir into national treatment regimens. Burundi approved the inclusion of TLD in the national guidelines for the use of antiretroviral drugs for the prevention and treatment of HIV as first-line treatment for men, male children over 35 kg, and women over 49 years of age (who are no longer of childbearing age). Pending sufficient data on the safety of Dolutegravir in women of childbearing age and in order to rule out any risk of fetal toxicity, the Government of Burundi has taken the interim decision not to authorize the use of TLD by female children and women of childbearing age. TLE [Tenefovir (TDF)/Lamivudine (3TC)/Efavirenz (EFV)] remains the first-line treatment regimen in young girls, women of childbearing age, pregnant women and lactating women.” | Yes | 2020 [2] |
| Kenya | Guidelines on the use of antiretroviral drugs for treating and preventing HIV infection in Kenya (2018) [3] | “DTG is preferred in first line ART in combination with two other ARVs for adolescents and adults. DTG is not recommended for women and adolescent girls of childbearing potential. Women and adolescent girls who are on effective contraception may opt to use DTG and should be supported in their decision.” | Yes | 2019 [4] |
| Uganda | Consolidated Guidelines for Prevention and Treatment of HIV in Uganda (2018) [5] | “The following categories of HIV positive patients are eligible for DTG-based regimen: All men and boys ≥ 6 years and women and adolescent girls with effective contraception or not of child-bearing potential. Note: Women of child-bearing potential not on contraceptives should be given information and counseled about the potential benefits and risks of DTG, including the risk of potential birth defects to allow for an informed decision on their ART regimen and contraceptive choices. If they choose DTG, their choice should be clearly documented and endorsed by the patient, parent or legal guardian in writing.” | Yes | 2020 [6] |
| Rwanda | Circular on changes in HIV prevention and management guidelines (2018) [7] | “Dolutegravir (DTG) (50mg)-based regimen is recommended for all newly enrolled adolescents and adults who weigh more than 35kgs except all female clients who are less than 50 years old.” | Yes | 2020 [8] |
| Cambodia | Operational Guidance: Use of Dolutegravir (DTG) for adults and adolescents in Cambodia (2018) [9] | “Effective January 2019, the new preferred first-line regimen for treatment-naive adults and adolescents (> 10 years and > 30kg) is now changed from TDF/3TC/EFV to TLD (TDF/3TC/DTG)…. Adolescent girls and women of childbearing potential who do not currently want to become pregnant can receive DTG together with consistent and reliable contraception. An EFV-based regimen is recommended for women desiring pregnancy or women with childbearing potential who do not wish to take contraception.” | Yes | 2021 [10] |
| Democratic Republic of Congo | Guide de prise en charge integrée du VIH en République Démocratique du Congo (2019) [11] | *Translation:* DTG-based regimens are recommended as first-line treatment for “adult and adolescent men from 30 kg”… and for “women of reproductive age who are using long-acting contraceptive methods, menopausal women, women unable to conceive and pregnant women after the first trimester of pregnancy.” | Yes | 2021 [12] |
| Indonesia | Rekomendasi Panel Ahli Penanggulangan HIV/AIDS dan IMS Hasil Rapat Daring, Kamis 2 Juli 2020. (2020) [13] | *Translation*: DTG recommended as first-line ART for all adolescent and adult patients starting treatment, “except women planning pregnancy and pregnant women in the first trimester,” for whom TDF+3TC+EFV should be used …“because there is not enough clinical evidence for the use of DTG in the 1st trimester.” | Yes | 2022 [14] |
| Cameroon | Directives nationales sur la prise en charge du VIH (2021) [15] | *Translation*: “Dolutegravir is preferred first-line treatment for male adults and adolescents and females above reproductive age. For females of reproductive age, DTG is preferred treatment regimen if contraceptive is being used, or if women not on contraception provide informed consent.” DTG listed among molecules that are contraindicated during pregnancy. | Yes | 2024 [16] |
| Congo | Lignes directrices relatives à l’utilisation de médicaments antirétroviraux pour le traitement et la prévention de l’infection à VIH au Congo (2019) [17] | *Translation*: “A regimen combining Dolutegravir is recommended as first-line treatment in adults, adolescents and children from 6 years of age starting antiretroviral treatment and in the event of failure of first-line treatment when the previous regimen did not contain Dolutegravir.” | No |  |
| Tanzania | National Guidelines for Management of HIV and AIDS (2019) [18] | “Dolutegravir-based regimen (TDF + 3TC + DTG [TLD]) is recommended as the preferred default first-line regimen for adults living with HIV. TLE will remain as an alternative for clients who will not tolerate and women at child-bearing potential who will not opt to use DTG. A women-centered approach is adopted and women of child bearing potential including those who are using long term effective contraception will be given adequate information to enable them to make informed decision and informed choice consent to using DTG.” | No |  |
| Vietnam | Guidelines for HIV/AIDS Treatment and Care (2019) [19] | “Use DTG for women and adolescent girls of childbearing age:  - Advise on the use of safe contraceptive methods and the possible risk of neural tube defects when using DTG in the first 3 months of pregnancy.  - DTG can still be prescribed in cases where regular, safe contraception is not used and patients still choose DTG after being fully counseled.” | No |  |
| India | National Guidelines for HIV Care and Treatment (2021) [20] | “NACO Technical Resource Group approved its usage since July 2020. NACO recommends Dolutegravir as the preferred drug for treatment of HIV-positive adults, adolescents and children (aged more than 6 years with bodyweight more than 20 kg) under the NACP.” | No |  |
| Malaysia | Malaysian Consensus Guidelines on Antiretroviral Therapy (2022) [21] | “The risk–benefit models suggest that the benefits of DTG for women of childbearing potential newly initiating ART, which include greater maternal viral suppression, fewer maternal deaths, fewer sexual transmissions and fewer mother-to-child transmissions, are likely to outweigh the risks.” | No |  |
| Thailand | Thailand National Guidelines on HIV/AIDS Treatment and Prevention (2021) [22] | *Translation*: “Recommended as first-line regimen: (TAF or TDF) + (3TC or FTC) + DTG. In cases where DTG is not yet available in the hospital or those infected cannot take DTG, consider choosing EFV or RPV instead. HIV regimens recommended as an alternative regimen: ABC + 3TC or AZT + 3TC plus DTG or EFV or RPV.” Recommendations for pregnant women: (TDF or TAF) + (3TC or FTC) +DTG, recommended as a combination drug (TDF/3TC/DTG) or (TAF/3TC/DTG). “Because the benefits of DTG are [exceed risks], the use of DTG is not prohibited in women of reproductive age.” | No |  |
| Philippines | Tenofovir/Lamivudine/Dolutegravir for treatment-naive and treatment-experienced adolescents and adults living with HIV (2021) [23] | “The National AIDS and STI Prevention and Control Program (NASPCP) of the DOH-DPCB proposed in the local implementing guidelines for treatment of PLHIV the shift to DTG-based regimens as (1) first line treatment for treatment-naive adolescents and adults living with HIV with TB co-infection; and; as (2) second-line treatment for treatment-experienced adults and adolescents living with HIV, specifically on those failing in TDF-based regimens.” | No |  |

11.	Ministère de la Santé Publique (République Démocratique du Congo). Guide de prise en charge integrée du VIH en République Démocratique du Congo (draft). . 2019.

12.	Ministère de la Santé Publique (République Démocratique du Congo). Guide de prise en charge integrée du VIH en République Démocratique du Congo. 2021.

13.	Expert Panel for HIV/AIDS and STI Prevention (Indonesia). Rekomendasi Panel Ahli Penanggulangan HIV/AIDS dan IMS Hasil Rapat Daring, Kamis 2 Juli 2020 2020 [October 1, 2024]. Available from: <https://hivaids-pimsindonesia.or.id/>.

14.	Ministry of Health (Indonesia). Regulation on Management of HIV, AIDS and STIs 2022 [September 25, 2024]. Available from: <https://peraturan.bpk.go.id/Details/245543/permenkes-no-23-tahun-2022>

15.	Ministere de la Santé Publique (Cameroon). Directives Nationales sur la Prise en Charge du VIH 2019 [September 24, 2024]. Available from: <https://hivpreventioncoalition.unaids.org/en/resources/national-guidelines-hiv-care>.

16.	(Cameroon) MoPH. Guide de Prévention et de Prise en Charge du VIH/SIDA au Cameroun. 2024.

17.	Ministere de la Sante et de la Population (Republic of Congo). Lignes directrices relatives à l’utilisation de médicaments antirétroviraux pour le traitement et la prévention de l’infection à VIH au Congo 2019.

18.	Ministry of Health (Tanzania). National Guidelines for the Management of HIV and AIDS. 2019 [September 24, 2024]. Available from: <https://hivpreventioncoalition.unaids.org/sites/default/files/attachments/national_guidelines_for_the_management_of_hiv_and_aids_2019.pdf>.

19.	Ministry of Health (Vietnam). Guidelines for HIV/AIDS Treatment and Care Issued together with Decision No. 5456/QD-BYT. 2019 [September 25, 2024].

20.	Ministry of Health and Family Welfare (India). National Guidelines for HIV Care and Treatment 2021 [September 24, 2024]. Available from: <https://naco.gov.in/sites/default/files/2020-21.pdf>.

21.	Malaysian Society for HIV Medicine (MASHM). Malaysian Consensus Guidelines on Antiretroviral Therapy 2022 [September 25, 2022]. Available from: <https://www.mashm.net/guidelines>.

22.	Department of Disease Control (Thailand). Thailand National Guidelines on HIV/AIDS Treatment and Prevention 2021 [September 25, 2024]. Available from: <https://www.thaiaidssociety.org/wp-content/uploads/2023/03/HIV-AIDS-Guideline-2564_2565_ED2.pdf>.

23.	Department of Health (Philippines). Tenofovir/Lamivudine/Dolutegravir for treatment-naive and treatment-experienced adolescents and adults living with HIV 2021 [September 25, 2024]. Available from: <https://hta.dost.gov.ph/wp-content/uploads/2021/09/ES_TLD_for_HIV_approved_by_SOH.docx.pdf>.

**Supplementary Table 2. Antiretroviral Therapy Regimens and Dolutegravir Use, by patient characteristic, sex and age group**

| **Characteristic** | **All Patients** | **Females**  **Aged 16-49 y** | **Males**  **Aged 16-49 y** | **Females**  **Aged ≥50 y** | **Males**  **Aged ≥50 y** |
| --- | --- | --- | --- | --- | --- |
| **Initial regimen base among patients newly initiating ART, n (%)** | **N=42380** | **N=23281** | **N=13415** | **N=2918** | **N=3281** |
| Dolutegravir | 24051 (56.8) | 10634 (45.7) | 9485 (70.7) | 1990 (68.2) | 1942 (70.2) |
| Efavirenz | 17663 (41.7) | 12278 (52.7) | 3784 (28.2) | 848 (29.1) | 753 (27.2) |
| Nevirapine | 404 (1) | 226 (1.0) | 82 (0.6) | 53 (1.8) | 43 (1.6) |
| Protease inhibitor | 243 (0.6) | 137 (0.6) | 59 (0.4) | 23 (0.8) | 24 (0.9) |
| Other* | 19 (0) | 6 (0) | 5 (0) | 4 (0.1) | 4 (0.1) |
| ART-naive patients who initiated a non-DTG regimen and later switched to dolutegravir, n (%) | 9167 (50.0) | 6100 (48.2) | 2026 (51.6) | 572 (61.6) | 469 (56.9) |
| **Latest regimen bases among all patients alive and engaged in care at database closure, n (%)** | **N=134284** | **N=67686** | **N=32530** | **N=19216** | **N=14852** |
| Dolutegravir | 103204 (76.9) | 49369 (72.9) | 26794 (82.4) | 15352 (79.9) | 11689 (78.7) |
| Not on dolutegravir | 31080 (23.1) | 18317 (27.1) | 5736 (17.6) | 3864 (20.1) | 3163 (21.3) |
| **Latest regimen bases among patients on non-DTG regimen at database closure, n (%)** | **N=31080** | **N=18317** | **N=5736** | **N=3864** | **N=3163** |
| Efavirenz | 16598 (53.4) | 11130 (60.8) | 2680 (46.7) | 1551 (40.1) | 1237 (39.1) |
| Nevirapine | 2564 (8.2) | 1185 (6.5) | 490 (8.5) | 508 (13.1) | 381 (12.0) |
| Protease inhibitor | 11662 (37.5) | 5916 (32.3) | 2478 (43.2) | 1775 (45.9) | 1493 (47.2) |
| Other* | 256 (0.8) | 86 (0.5) | 88 (1.5) | 30 (0.8) | 52 (1.6) |

*Other regimen bases include other integrase inhibitors (Bictegravir, Raltegravir, Elvitegravir) and NNRTIs (Etravirine)

**Supplementary Figure 2. Regimen base among patients not on DTG-based ART at database closure (July 31, 2021) across all sites (n=31,080)**

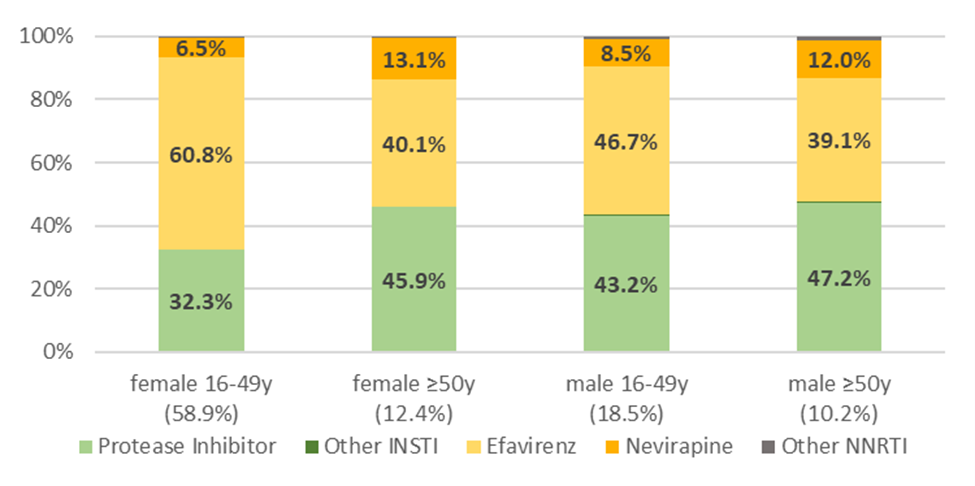

**Supplementary Table 3. Cumulative Incidence of Dolutegravir Uptake, by Sex and Age Group, excluding clinics in Kenya**

| **Variable** | **Summary of Outcomes** | | | | **Dolutegravir Uptake, Cumulative Incidence Proportion (95% CI)** | | | |
| --- | --- | --- | --- | --- | --- | --- | --- | --- |
|  | **Total Patients, *n*** | **Patients Initiating Dolutegravir, *n*** | **Patients Who Dropped Out, *n*** | **Patients Censored, *n*** | **Before the Safety Signal**  **(Until 18 May 2018)** | **After the Safety Signal (Until 22 July 2019)** | **One year after WHO Recommended Dolutegravir for All (Until July 22 2020)** | **Two years after WHO Recommended Dolutegravir for All (Until July 22 2021)** |
| **Entire sample** | 91692 | 64366 | 12163 | 15163 | 1.3 (1.2, 1.3) | 21.7 (21.5, 22.0) | 42.7 (42.4, 43) | 69.9 (69.6, 70.2) |
| **Sex and age group** |  |  |  |  |  |  |  |  |
| Females aged 16-49 y | 46070 | 30449 | 6965 | 8656 | 1.2 (1.2, 1.4) | 10.9 (10.7, 11.2) | 29.5 (29.1, 29.9) | 65.8 (65.3, 66.2) |
| Males aged 16-49 y | 11865 | 8784 | 1202 | 1879 | 1.4 (1.3, 1.6) | 33.6 (33.0, 34.2) | 57.3 (56.7, 58.0) | 75.2 (74.7, 75.8) |
| Females aged ≥50 y | 23604 | 17808 | 2886 | 2910 | 1.0 (0.8, 1.1) | 29.9 (29.1, 30.8) | 53.4 (52.5, 54.3) | 73.8 (73.0, 74.6) |
| Males aged ≥50 y | 10153 | 7325 | 1110 | 1718 | 1.4 (1.2, 1.6) | 33.5 (32.6, 34.5) | 56.1 (55.2, 57.1) | 71.9 (71.0, 72.8) |

**Supplementary Table 4. Sample of patients at sites with a database closure after July 31, 2022 (63 clinics in 8 countries)**

| **Characteristic** | **All Patients** | **Females**  **Aged 16-49 y** | **Males**  **Aged 16-49 y** | **Females**  **Aged ≥50 y** | **Males**  **Aged ≥50 y** |
| --- | --- | --- | --- | --- | --- |
| Total, n (%) | 182824 | 94804 (51.9) | 45575 (24.9) | 23772 (13.0) | 18673 (10.2) |
| Mean age (SD), y | 41.3 (11.4) | 35.8 (8.0) | 38.1 (7.3) | 56.7 (5.9) | 57.2 (6.2) |
| **Experience with ART at start of follow-up, n (%)** | | | | | |
| Experienced | 132117 (72.3) | 66830 (70.5) | 20274 (85.3) | 29620 (65.0) | 15393 (82.4) |
| Naive | 50707 (27.7) | 27974 (29.5) | 3498 (14.7) | 15955 (35.0) | 3280 (17.6) |
| **Site urbanicity, *n* (*%*)** * |  |  |  |  |  |
| Rural | 38627 (21.1) | 19923 (21.0) | 5419 (22.8) | 9493 (20.8) | 3792 (20.3) |
| Urban | 144197 (78.9) | 74881 (79.0) | 18353 (77.2) | 36082 (79.2) | 14881 (79.7) |
| **Site level of care, *n* (*%*)** * |  |  |  |  |  |
| Primary | 51968 (28.4) | 28041 (29.6) | 6347 (26.7) | 12822 (28.1) | 4758 (25.5) |
| Secondary | 45463 (24.9) | 23906 (25.2) | 6034 (25.4) | 11246 (24.7) | 4277 (22.9) |
| Tertiary | 85393 (46.7) | 42857 (45.2) | 11391 (47.9) | 21507 (47.2) | 9638 (51.6) |
| **Country, *n* (*%*)** |  |  |  |  |  |
| Burundi | 6649 (3.6) | 3057 (3.2) | 1371 (5.8) | 1151 (2.5) | 1070 (5.7) |
| Cameroon | 13810 (7.6) | 6955 (7.3) | 2191 (9.2) | 3308 (7.3) | 1356 (7.3) |
| Congo | 6236 (3.4) | 3047 (3.2) | 1445 (6.1) | 757 (1.7) | 987 (5.3) |
| Democratic Republic of Congo | 4019 (2.2) | 2249 (2.4) | 603 (2.5) | 722 (1.6) | 445 (2.4) |
| Kenya | 90570 (49.5) | 47325 (49.9) | 11948 (50.3) | 22302 (48.9) | 8995 (48.2) |
| Rwanda | 18199 (10) | 9254 (9.8) | 1775 (7.5) | 5439 (11.9) | 1731 (9.3) |
| Tanzania | 10401 (5.7) | 5604 (5.9) | 1368 (5.8) | 2418 (5.3) | 1011 (5.4) |
| Uganda | 32940 (18) | 17313 (18.3) | 3071 (12.9) | 9478 (20.8) | 3078 (16.5) |
| **Initial guidelines on DTG-based regimens for 1st-line ART, *n* (*%*)** | | | | | |
| Restrictive guidelines for WRA† | 166187 (90.9) | 86153 (90.9) | 20959 (88.2) | 42400 (93) | 16675 (89.3) |
| Non-restrictive guidelines for WRA‡ | 16637 (9.1) | 8651 (9.1) | 2813 (11.8) | 3175 (7.0) | 1998 (10.7) |
| **Country income, *n* (*%*)** |  |  |  |  |  |
| Low income | 61807 (33.8) | 31873 (33.6) | 6820 (28.7) | 16790 (36.8) | 6324 (33.9) |
| Lower-middle income | 121017 (66.2) | 62931 (66.4) | 16952 (71.3) | 28785 (63.2) | 12349 (66.1) |
| **Region, *n* (*%*)** |  |  |  |  |  |
| Central Africa | 48913 (26.8) | 24562 (25.9) | 7385 (31.1) | 11377 (25) | 5589 (29.9) |
| East Africa | 133911 (73.2) | 70242 (74.1) | 16387 (68.9) | 34198 (75) | 13084 (70.1) |

ART: Antiretroviral therapy. DTG: Dolutegravir. SD: Standard deviation. WRA: Women of reproductive age.

† Burundi, Cameroon, Democratic Republic of Congo, Kenya, Rwanda, Uganda

‡ Congo, Tanzania

**Supplementary Table 5. Cumulative Incidence of Dolutegravir Uptake by July 31, 2022, by patient characteristics**

|  | **Summary of Outcomes** | | | | **Dolutegravir Uptake, Cumulative Incidence Proportion (95% CI)** | | | | |
| --- | --- | --- | --- | --- | --- | --- | --- | --- | --- |
| **Characteristic** | **Total Patients, *n*** | **Patients Initiating Dolutegravir, *n*** | **Patients Who Dropped Out, *n*** | **Patients Censored, *n*** | **Before the Safety Signal (Until 18 May 2018)** | **After the Safety Signal (Until 22 July 2019)** | **One year after WHO Recommended Dolutegravir for All (Until July 22 2020)** | **Two years after  WHO Recommended Dolutegravir for All (Until July 22 2021)** | **Three years after WHO Recommended Dolutegravir for All (Until July 22 2022)** |
| **Entire sample** | 182824 | 144140 | 25300 | 13384 | 2.7 (2.6, 2.7) | 28.7 (28.5, 28.9) | 46.2 (46, 46.5) | 68.9 (68.7, 69.1) | 78.7 (78.5, 78.9) |
| **Sex and age group** |  |  |  |  |  |  |  |  |  |
| Females aged 16-49 y | 94804 | 71871 | 15444 | 7489 | 2.5 (2.4, 2.6) | 15.5 (15.3, 15.7) | 32.1 (31.8, 32.4) | 63.5 (63.2, 63.8) | 75.7 (75.4, 75.9) |
| Males aged 16-49 y | 23772 | 19546 | 2313 | 1913 | 2.2 (2, 2.3) | 39.8 (39.4, 40.3) | 59.6 (59.1, 60.0) | 73.7 (73.3, 74.1) | 82.1 (81.8, 82.5) |
| Females aged ≥50 y | 45575 | 37472 | 5601 | 2502 | 3.7 (3.4, 3.9) | 45.4 (44.8, 46.1) | 63.1 (62.5, 63.7) | 76.3 (75.7, 76.8) | 82.2 (81.7, 82.6) |
| Males aged ≥50 y | 18673 | 15251 | 1942 | 1480 | 3.4 (3.1, 3.6) | 47.0 (46.3, 47.7) | 64.2 (63.5, 64.9) | 75.4 (74.8, 76.0) | 81.6 (81.1, 82.2) |
| **Experience with ART at start of follow-up** | | |  |  |  |  |  |  |  |
| **Naïve** |  |  |  |  |  |  |  |  |  |
| Females aged 16-49 y | 27974 | 22309 | 4999 | 666 | 0.9 (0.8, 1.1) | 6.2 (5.9, 6.5) | 25.0 (24.5, 25.5) | 59.5 (58.9, 60.1) | 79.4 (79.0, 79.9) |
| Males aged 16-49 y | 15955 | 14192 | 1559 | 204 | 1.0 (0.9, 1.2) | 22.3 (21.6, 22.9) | 48.1 (47.3, 48.8) | 71.5 (70.8, 72.2) | 88.7 (88.2, 89.2) |
| Females aged ≥50 y | 3498 | 3170 | 265 | 63 | 0.9 (0.6, 1.2) | 21.8 (20.4, 23.2) | 46.2 (44.5, 47.8) | 72.9 (71.4, 74.3) | 90.5 (89.4, 91.4) |
| Males aged ≥50 y | 3280 | 2961 | 266 | 53 | 1.1 (0.8, 1.5) | 23.4 (22.0, 24.9) | 47.6 (45.8, 49.3) | 72.2 (70.6, 73.7) | 90.1 (89.0, 91.0) |
| **Experienced** |  |  |  |  |  |  |  |  |  |
| Females aged 16-49 y | 66830 | 49562 | 10445 | 6823 | 3.2 (3.1, 3.3) | 19.4 (19.1, 19.7) | 35.0 (34.7, 35.4) | 65.2 (64.8, 65.5) | 74.1 (73.7, 74.4) |
| Males aged 16-49 y | 29620 | 23280 | 4042 | 2298 | 2.8 (2.6, 3.0) | 49.3 (48.7, 49.9) | 65.8 (65.2, 66.3) | 74.9 (74.4, 75.4) | 78.6 (78.1, 79.0) |
| Females aged ≥50 y | 20274 | 16376 | 2048 | 1850 | 4.1 (3.9, 4.4) | 49.5 (48.8, 50.2) | 66.1 (65.4, 66.7) | 76.9 (76.3, 77.4) | 80.7 (80.2, 81.3) |
| Males aged ≥50 y | 15393 | 12290 | 1676 | 1427 | 3.8 (3.5, 4.1) | 52.1 (51.3, 52.8) | 67.7 (67.0, 68.5) | 76.1 (75.4, 76.8) | 79.8 (79.2, 80.4) |

**Supplementary Table 6. Cumulative Incidence of Dolutegravir Uptake by July 31, 2022, by contextual characteristics**

|  | **Summary of Outcomes** | | | | **Dolutegravir Uptake, Cumulative Incidence Proportion (95% CI)** | | | | |
| --- | --- | --- | --- | --- | --- | --- | --- | --- | --- |
| **Characteristic** | **Total Patients, *n*** | **Patients Initiating Dolutegravir, *n*** | **Patients Who Dropped Out, *n*** | **Patients Censored, *n*** | **Before the Safety Signal (Until 18 May 2018)** | **After the Safety Signal (Until 22 July 2019)** | **One year after WHO Recommended Dolutegravir for All (Until July 22 2020)** | **Two years after  WHO Recommended Dolutegravir for All (Until July 22 2021)** | **Three years after WHO Recommended Dolutegravir for All (Until July 22 2022)** |
| **Initial guidelines on DTG for first-line ART** | | |  |  |  |  |  |  |  |
| **Restriction or strong caution for females of reproductive age†** | **166187** | **130611** | **24073** | **11503** | **2.9 (2.9-3.0)** | **29.5 (29.3, 29.7)** | **45.9 (45.7, 46.1)** | **68.6 (68.4, 68.8)** | **78.5 (78.3, 78.7)** |
| Females aged 16-49 y | 86153 | 64832 | 14869 | 6452 | 2.8 (2.7-2.9) | 15.2 (14.9, 15.4) | 30.2 (29.9, 30.5) | 62.7 (62.3, 63) | 75.1 (74.8, 75.4) |
| Males aged 16-49 y | 42400 | 34733 | 5392 | 2275 | 2.3 (2.2-2.5) | 41.3 (40.8, 41.8) | 59.8 (59.3, 60.2) | 73.5 (73.1, 73.9) | 81.8 (81.5, 82.2) |
| Females aged ≥50 y | 20959 | 17327 | 2082 | 1550 | 4.2 (3.9-4.4) | 48.4 (47.7, 49.0) | 65.8 (65.2, 66.5) | 77.1 (76.5, 77.6) | 82.6 (82.1, 83.1) |
| Males aged ≥50 y | 16675 | 13719 | 1730 | 1226 | 3.8 (3.5-4.0) | 49.9 (49.1, 50.6) | 66.5 (65.8, 67.3) | 76.4 (75.7, 77.0) | 82.2 (81.6, 82.8) |
| **No restriction or strong caution for females of reproductive age‡** | **16637** | **13529** | **1227** | **1881** | **NA** | **20.6 (20.0, 21.2)** | **49.7 (48.9, 50.5)** | **71.8 (71.2, 72.5)** | **81.2 (80.6, 81.8)** |
| Females aged 16-49 y | 8651 | 7039 | 575 | 1037 | NA | 19.0 (18.2, 19.9) | 50.4 (49.4, 51.5) | 71.8 (70.8, 72.7) | 81.2 (80.4, 82.0) |
| Males aged 16-49 y | 3175 | 2739 | 209 | 227 | NA | 20.3 (18.9, 21.7) | 56.8 (55.0, 58.5) | 76.1 (74.6, 77.6) | 86.2 (84.9, 87.3) |
| Females aged ≥50 y | 2813 | 2219 | 231 | 363 | NA | 23.6 (22.1, 25.2) | 43.2 (41.3, 45.0) | 70.6 (68.8, 72.2) | 78.8 (77.2, 80.2) |
| Males aged ≥50 y | 1998 | 1532 | 212 | 254 | NA | 23.4 (21.5, 25.3) | 44.5 (42.4, 46.7) | 67.2 (65.1, 69.2) | 76.5 (74.6, 78.3) |
| **Country income level** |  |  |  |  |  |  |  |  |  |
| **Low-income** | **61807** | **52254** | **7135** | **2418** | **1.8 (1.6, 1.9)** | **26.2 (25.9, 26.6)** | **46**.0 **(45.6, 46.4)** | **73**.0 **(72.6, 73.3)** | **84.5 (84.2, 84.8)** |
| Females aged 16-49 y | 31873 | 25958 | 4584 | 1331 | 1.8 (1.6, 1.9) | 10.4 (10.1, 10.8) | 26.1 (25.6, 26.6) | 66.2 (65.7, 66.7) | 81.3 (80.9, 81.8) |
| Males aged 16-49 y | 16790 | 14781 | 1571 | 438 | 1.7 (1.5, 1.9) | 42.6 (41.9, 43.4) | 65.2 (64.5, 65.9) | 79.5 (78.9, 80.1) | 88.0 (87.5, 88.4) |
| Females aged ≥50 y | 6820 | 5998 | 500 | 322 | 1.6 (1.4, 2.0) | 42.1 (40.9, 43.3) | 69.4 (68.3, 70.5) | 81.6 (80.6, 82.5) | 88.0 (87.2, 88.7) |
| Males aged ≥50 y | 6324 | 5517 | 480 | 327 | 1.9 (1.6, 2.2) | 45.4 (44.1, 46.6) | 70.0 (68.9, 71.1) | 80.5 (79.5, 81.5) | 87.2 (86.4, 88.0) |
| **Lower-middle income** | **121017** | **91886** | **18165** | **10966** | **3.1 (3**.0**, 3.2)** | **29.9 (29.7, 30.2)** | **46.4 (46.1, 46.7)** | **66.8 (66.6, 67.1)** | **75.8 (75.5, 76**.0**)** |
| Females aged 16-49 y | 62931 | 45913 | 10860 | 6158 | 2.9 (2.8, 3.1) | 18.1 (17.8, 18.4) | 35.1 (34.7, 35.5) | 62.1 (61.7, 62.5) | 72.8 (72.4, 73.1) |
| Males aged 16-49 y | 28785 | 22691 | 4030 | 2064 | 2.4 (2.2, 2.6) | 38.2 (37.6, 38.8) | 56.3 (55.7, 56.8) | 70.3 (69.8, 70.8) | 78.7 (78.2, 79.2) |
| Females aged ≥50 y | 16952 | 13548 | 1813 | 1591 | 4.5 (4.2, 4.8) | 46.8 (46.0, 47.5) | 60.6 (59.9, 61.4) | 74.2 (73.5, 74.8) | 79.8 (79.2, 80.4) |
| Males aged ≥50 y | 12349 | 9734 | 1462 | 1153 | 4.1 (3.8, 4.5) | 47.9 (47.0, 48.7) | 61.2 (60.4, 62.1) | 72.8 (72.0, 73.6) | 78.7 (78.0, 79.5) |
| **Health system level** |  |  |  |  |  |  |  |  |  |
| **Health center** | **51968** | **41626** | **6899** | **3443** | **1.2 (1.1, 1.3)** | **27.7 (27.3, 28.1)** | **50.7 (50.3, 51.1)** | **69.5 (69.1, 69.9)** | **80.0 (79.6, 80.3)** |
| Females aged 16-49 y | 28041 | 21517 | 4509 | 2015 | 1.1 (1.0, 1.2) | 13.1 (12.7, 13.4) | 35.3 (34.8, 35.9) | 62.7 (62.1, 63.2) | 76.6 (76.1, 77.1) |
| Males aged 16-49 y | 12822 | 10725 | 1485 | 612 | 0.8 (0.7, 1.0) | 41.0 (40.1, 41.8) | 64.9 (64.1, 65.7) | 75.6 (74.8, 76.3) | 83.5 (82.9, 84.2) |
| Females aged ≥50 y | 6347 | 5388 | 492 | 467 | 2.1 (1.7, 2.4) | 49.0 (47.8, 50.3) | 73.5 (72.4, 74.6) | 80.3 (79.3, 81.3) | 84.8 (83.9, 85.7) |
| Males aged ≥50 y | 4758 | 3996 | 413 | 349 | 1.7 (1.4, 2.1) | 49.7 (48.3, 51.1) | 72.5 (71.2, 73.7) | 79.3 (78.1, 80.4) | 84.0 (82.9, 85.0) |
| **District hospital** | **45463** | **34927** | **7133** | **3403** | **3.7 (3.5, 3.9)** | **33.7 (33.2, 34.1)** | **48.1 (47.7, 48.6)** | **67.4 (66.9, 67.8)** | **76.7 (76.3, 77.0)** |
| Females aged 16-49 y | 23906 | 17408 | 4607 | 1891 | 3.5 (3.2, 3.7) | 18.0 (17.5, 18.4) | 31.6 (31.0, 32.2) | 60.3 (59.7, 60.9) | 72.6 (72.0, 73.1) |
| Males aged 16-49 y | 11246 | 8992 | 1551 | 703 | 2.5 (2.2, 2.8) | 43.1 (42.2, 44.0) | 61.0 (60.1, 61.9) | 72.1 (71.3, 73.0) | 79.9 (79.1, 80.6) |
| Females aged ≥50 y | 6034 | 5027 | 546 | 461 | 6.1 (5.5, 6.7) | 60.0 (58.8, 61.3) | 73.2 (72.1, 74.3) | 79.7 (78.6, 80.6) | 83.2 (82.3, 84.2) |
| Males aged ≥50 y | 4277 | 3500 | 429 | 348 | 4.6 (4, 5.3) | 59.4 (57.9, 60.8) | 71.0 (69.7, 72.4) | 77.2 (75.9, 78.4) | 81.8 (80.6, 82.9) |
| **Regional, provincial, university hospital** | **85393** | **67587** | **11268** | **6538** | **3 (2.9, 3.1)** | **26.6 (26.3, 26.9)** | **42.5 (42.2, 42.9)** | **69.4 (69.1, 69.7)** | **79.0 (78.8, 79.3)** |
| Females aged 16-49 y | 42857 | 32946 | 6328 | 3583 | 3 (2.8, 3.1) | 15.7 (15.4, 16.1) | 30.2 (29.7, 30.6) | 65.8 (65.4, 66.3) | 76.8 (76.4, 77.2) |
| Males aged 16-49 y | 21507 | 17755 | 2565 | 1187 | 2.8 (2.6, 3) | 37.4 (36.8, 38.1) | 55.6 (54.9, 56.3) | 73.4 (72.8, 74.0) | 82.4 (81.9, 82.9) |
| Females aged ≥50 y | 11391 | 9131 | 1275 | 985 | 3.3 (3, 3.6) | 35.7 (34.8, 36.6) | 52.0 (51.1, 52.9) | 72.3 (71.4, 73.1) | 80.1 (79.4, 80.8) |
| Males aged ≥50 y | 9638 | 7755 | 1100 | 783 | 3.6 (3.2, 4) | 40.2 (39.2, 41.2) | 57.1 (56.1, 58.1) | 72.7 (71.8, 73.6) | 80.4 (79.6, 81.2) |
